# A Novel Composite Index for Cardiovascular Risk: The Cholesterol, high-density lipoprotein, Glucose (CHG) Index

**DOI:** 10.1101/2025.04.02.25325143

**Authors:** Amin Mansoori, Mohadeseh Poudineh, Maryam Dianati, Gordon Ferns, Mark Ghamsary, Habibollah Esmaily, Mohsen Moohebati, Majid Ghayour-Mobarhan

**Affiliations:** Department of Applied Mathematics, School of Mathematical Sciences, Ferdowsi University of Mashhad, Mashhad, Iran; School of Medicine, Zanjan University of Medical Sciences, Zanjan, Iran; Student Research Committee, Rafsanjan University of Medical Sciences, Rafsanjan, Iran; Brighton and Sussex Medical School, Division of Medical Education, Brighton, United Kingdom; School of public health, Department of Epidemiology and Biostatistics, Loma Linda University, Loma Linda, USA; Department of Biostatistics, School of Health, Mashhad University of Medical Sciences, Mashhad, Iran; Social Determinants of Health Research Center, Mashhad University of Medical Sciences, Mashhad, Iran; Cardiovascular Research Center, School of Medicine, Mashhad University of Medical Sciences, Mashhad, Iran; International UNESCO center for Health-Related Basic Sciences and Human Nutrition, Mashhad University of Medical Sciences, Mashhad, Iran

**Keywords:** CHG, TyG, Cardiovascular diseases

## Abstract

**Background:** Cardiovascular diseases (CVDs) are a leading cause of death, with traditional risk factors like dyslipidemia, insulin resistance (IR), and obesity studied extensively. However, current indices do not fully integrate glucose and cholesterol metabolism. This study introduces the Cholesterol, HDL, and Glucose Index (CHG) as a more comprehensive biomarker for diagnosing CVDs and compares its predictive value to existing indices like TyG, AIP, and LAP.

**Materials and Methods:** The Mashhad Stroke and Heart Atherosclerotic Disorder (MASHAD) study is a longitudinal prospective cohort study initiated in 2010, involving 7,641 participants. Over a 10-year follow-up, expert cardiologists diagnosed CVDs based on clinical evaluations and medical history. Receiver operating characteristic (ROC) curve analysis was employed to determine the cut-off values of CHG and other indices such as TyG, AIP, and LAP and compare their sensitivity and specificity.

**Results:** Among the 7,433 participants, 837 were diagnosed with CVDs. CHG showed the highest discriminatory power in assessing CVDs risk, with an area under the curve (AUC) of 0.660 (95% CI: 0.649–0.671, p<0.001) and a cut-off value of 5.28. TyG, LAP, and AIP showed lower AUC values of 0.637 (95% CI: 0.626–0.648, p<0.001), 0.599 (95% CI: 0.588–0.611, p<0.001), and 0.592 (95% CI: 0.580–0.603, p<0.001), respectively, with corresponding cut-off values of 8.93, 40.42, and 0.38.

**Conclusion:** CHG is an innovative composite index that combines glucose and lipid markers, enhancing its predictive capability for diagnosing cardiovascular diseases. This index delivers a more thorough evaluation of cardiometabolic health, taking into account the intricate relationship between glucose and lipid metabolism in the development of CVDs.

## 1. Introduction

Cardiovascular diseases (CVDs) are the leading cause of death globally. Traditional risk factors such as dyslipidemia, insulin resistance (IR), and obesity have been extensively studied (1). Despite advancements in risk stratification, there remains a need for more comprehensive biomarkers that integrate multiple metabolic disturbances to enhance early detection and intervention.

IR occurs when tissues respond poorly to insulin, which hampers glucose uptake, oxidation, and glycogen synthesis, while also reducing the ability to suppress lipid oxidation. Studies indicate that although free fatty acids are the main energy source for ATP production in the adult heart, the heart's metabolic system is flexible and can use other substrates like glucose, lactate, and amino acids (2). However, during IR, these metabolic issues can lead to CVDs. A significant effect of IR is the disruption of glucose metabolism, which causes chronic high blood sugar levels. This condition promotes oxidative stress and inflammation, leading to cellular damage (3). Moreover, IR affects lipid metabolism, resulting in dyslipidemia, which is marked by the “lipid triad": (1) increased plasma triglycerides, (2) lower levels of high-density lipoprotein (HDL), and (3) a rise in small, dense low-density lipoproteins (LDL). Together with endothelial dysfunction—another result of impaired insulin signaling—these elements speed up the formation of atherosclerotic plaques, further heightening the risk of CVDs (1).

Several metabolic indices, such as the Triglyceride-Glucose Index (TyG), Atherogenic Index of Plasma (AIP), and Lipid Accumulation Product (LAP), are commonly used to evaluate CVDs and metabolic risk (4). However, these indices tend to focus either on lipid metabolism or insulin resistance, failing to fully consider the interaction between glucose and cholesterol metabolism. For example, while the TyG index is a dependable marker of insulin resistance, it does not address the atherogenic risk associated with cholesterol (5). Likewise, AIP and LAP highlight lipid abnormalities but neglect the role of glucose dysregulation in metabolic syndrome and CVDs (6). Due to these limitations, there is an increasing demand for an index that integrates both glucose and lipid markers to offer a more thorough evaluation of cardiometabolic health.

Given the intricate interplay between glucose and lipid metabolism in the pathogenesis of CVD, emerging composite biomarkers that incorporate both parameters may offer better predictive value than traditional lipid ratios. In this study, we aimed to introduce Cholesterol, HDL, and Glucose Index (CHG) as an alternative marker for diagnosing cardiovascular diseases (CVDs). We also assessed the predictive value of CHG in comparison to well-known indices TyG, LAP, and AIP.

## 2. Materials and Methods

### 2.1. Study Population

The Mashhad Stroke and Heart Atherosclerotic Disorder (MASHAD) study, launched in 2010, is a longitudinal prospective cohort study. In its first phase, a total of 9,704 individuals aged 35 to 65 were initially enrolled. After a decade of follow-up, phase II commenced with a reduced cohort of approximately 7,641 participants. Among them, 952 individuals were diagnosed with CVD by expert cardiologists. All participants provided written informed consent before enrolling in the study. The study protocol was thoroughly reviewed and received approval from the Ethics Committee of Mashhad University of Medical Sciences under the approval number IR.MUMS.REC.1386.250 (approved in July 2007).

Throughout the 10-year follow-up period, an experienced cardiologist conducted physical examinations and assessed participants' medical histories to identify CVD cases. The diagnosis was confirmed based on a history of myocardial infarction, angina pectoris, or the presence of a Q wave on an electrocardiogram. In cases where the diagnosis remained uncertain, additional tests, including exercise tolerance testing (ETT), echocardiography, stress echocardiography, and radioisotope imaging, were performed to ensure accuracy.

### 2.2. Data Collection

After a 14-hour overnight fast, blood samples were collected through venipuncture of a peripheral vein. Serum levels of triglycerides (TG), low-density lipoprotein cholesterol (LDL-C), high-density lipoprotein cholesterol (HDL-C), and total cholesterol (TC) were measured using previously established methods (7).

Height, weight, body mass index (BMI), waist circumference (WC), hip circumference (HC), waist-to-hip ratio (WHR), and mid-upper arm circumference (MAC) were assessed following standardized protocols for all participants, as previously described (7). Height, WC, HC, and MAC were measured to the nearest millimeter using a tape measure, while weight was recorded to the nearest 0.1 kg using electronic scales. BMI was determined by dividing weight (kg) by height squared (m^2^) (7). According to the World Health Organization (WHO), overweight is classified as a BMI between 25 and 29.9 kg/m^2^, while obesity is defined as a BMI of 30 kg/m^2^ or higher (8). WHR was calculated by dividing WC by HC. Based on the International Diabetes Federation (IDF) criteria, a WC of ≥80 cm for women and ≥94 cm for men is considered elevated (7).

The parameters used in the study are measured based on standard definitions and are presented in Table 1.

**Table 1.**
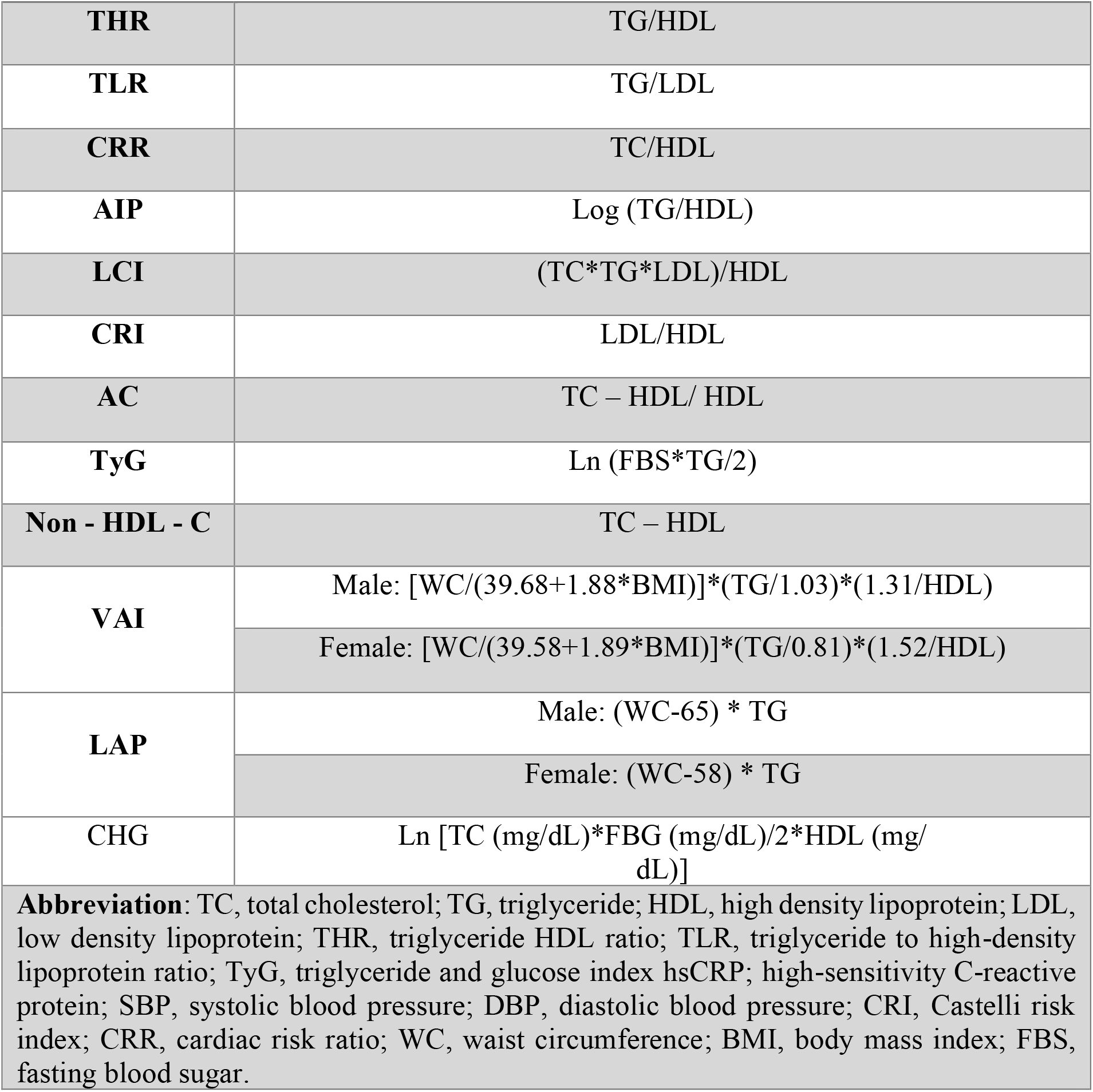
Calculation of parameters used in our study.

### 2.3. Statistical Analysis

All statistical analyses were conducted using SPSS version 20 (SPSS, Chicago, IL, USA) and MedCalc, version 22 (MedCalc Software, Ostend, Belgium). For normally distributed quantitative variables, the mean and standard deviation (SD) were reported. The Chi-square test was applied to compare qualitative variables. Receiver operating characteristic (ROC) curve analysis was utilized to determine the optimal cut-off values for TyG, TyG-BMI, TyG-WC, CHG, CHG-BMI, and CHG-WC, assessing their sensitivity and specificity in diagnosing type 2 diabetes mellitus. The Youden index was used to establish these thresholds. A p-value of less than 0.05 was considered statistically significant.

## 3. Result

### 3.1. Characteristics of the participants

Table 2 summarized the demographic and clinical characteristics of the study population. In this study, a total of 7,433 participants were included, with 837 individuals diagnosed with cardiovascular disease (CVD+) and 6,596 without CVD (CVD-). Participants with CVD significantly older than those without CVD, with a mean age of 51.30±7.50 years compared to 46.91±7.85 years (p<0.001). The proportion of females was higher in both groups, but a greater percentage was observed in the CVD-group (61.10%) compared to the CVD+ group (54.84%) (p<0.001).

**Table 2.** Clinical characteristics of the participants.

| **Table 2. Clinical characteristics of the participants** |  |  |  |
| --- | --- | --- | --- |
| **Variable** | **CVD+ (n=837)** | **CVD- (n=6596)** | **P-value\*** |
| **Age (year) (Mean±SD)** | 51.30±7.50 | 46.91±7.85 | <0.001 |
| **Sex (n)** |  |  |  |
| **Male** | 378 (45.16 %) | 2566 (38.90 %) | <0.001 |
| **Female** | 459 (54.84 %) | 4030 (61.10 %) |  |
| **TLR** | 1.69±3.01 | 1.36±1.78 | <0.001 |
| **CRR** | 4.94±1.24 | 4.56±1.11 | <0.001 |
| **LCI** | 0.20±0.14 | 0.17±0.11 | <0.001 |
| **nonHDL-C** | 157.24±40.31 | 146.74±35.98 | <0.001 |
| **AIP** | 0.54±0.27 | 0.45±0.27 | <0.001 |
| **CRI** | 2.98±1.01 | 2.79±0.91 | <0.001 |
| **AC** | 3.94±1.24 | 3.56±1.11 | <0.001 |
| **THR** | 4.30±3.55 | 3.47±2.69 | <0.001 |
| **TyG** | 8.86±0.71 | 8.52±0.62 | <0.001 |
| **VAI** | 3.18±2.92 | 2.57±1.98 | <0.001 |
| **LAP** | 69.58±55.40 | 54.78±43.97 | <0.001 |
| **CHG** | 5.48±0.46 | 5.23±0.37 | <0.001 |
| * Significant at a level of 0.05,<br>Values are reported as frequency (percent) and mean ± SD<br>**Abbreviations:** THR, triglyceride HDL ratio; TLR, triglyceride to high-density lipoprotein ratio; TyG, triglyceride and glucose index; CRI, castelli risk index; CRR, cardiac risk ratio; AC, atherogenic coefficient; AIP, atherogenic *index* of plasma; LCI, lipoprotein combined index; LAP, lipid accumulation product; VAI, visceral adiposity index; CHG, cholesterol, high-density lipoprotein, and Glucose index. |  |  |  |

**Table 3.**
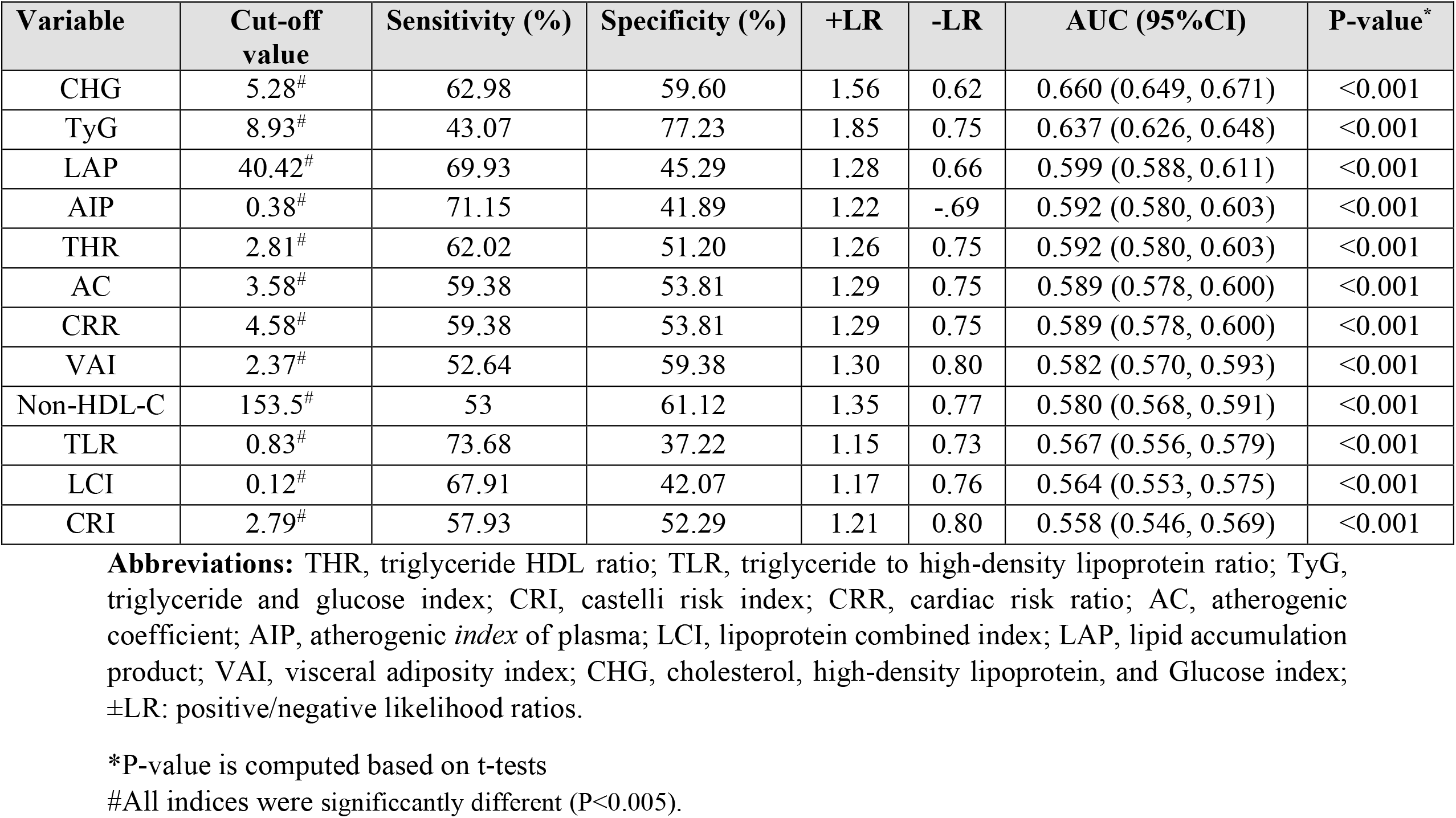
Areas under the curve (AUCs) and cut-off points for lipid profile of CVDs patients.

### 3.2. ROC Curves and estimation of cut-off points

CHG demonstrated the highest discriminatory power among the evaluated lipid and cardiometabolic indices for CVD, with an AUC of 0.660 (95% CI: 0.649–0.671, p<0.001) and a cut-off value of 5.28. Among other markers, TyG had an AUC of 0.637 (95% CI: 0.626–0.648, p<0.001) with a cut-off value of 8.93. LAP showed an AUC of 0.599 (95% CI: 0.588–0.611, p<0.001) with a cut-off of 40.42, while AIP had an AUC of 0.592 (95% CI: 0.580–0.603, p<0.001) with a cut-off of 0.38. These results highlight CHG as a promising novel biomarker with superior predictive value compared to traditional lipid indices in assessing CVD risk.

## 4. Discussion

CVDs remains a leading cause of morbidity and mortality worldwide, necessitating the identification of reliable and accessible biomarkers for early risk assessment. In this study, we investigated the predictive power of several lipid and cardiometabolic indices, with a particular focus on CHG as a novel biomarker.

Our findings revealed that CHG exhibited the highest area under the curve among all tested indices, outperforming traditional markers such as TyG, LAP, and AIP, likely due to their heavy reliance on triglyceride-based measurements, which may not fully capture the complex interplay between glucose metabolism and lipid abnormalities. The optimal cut-off value for CHG was 5.28, with a sensitivity of 62.98% and specificity of 59.60%, suggesting its potential utility in CVD risk stratification. Our previous study on patients with T2DM, the CHG index demonstrated the highest diagnostic performance among the evaluated indices, with an AUC of 0.864 (95% CI: 0.857– 0.871). At a cut-off value of 5.57, CHG exhibited a sensitivity of 70.38% and a specificity of 89.82% (9), reinforcing its potential as a robust biomarker for CVD risk assessment in this patient population.

The innovation of CHG is its ability to merge cholesterol, HDL, and glucose levels into a single metric. Unlike traditional lipid ratios that mainly concentrate on lipid metabolism (such as total cholesterol/HDL or triglycerides/HDL), CHG includes glucose levels, which are vital in understanding metabolic dysfunction and the progression of atherosclerosis (10, 11). This distinctive combination could provide a more thorough evaluation of cardiometabolic health by highlighting both lipid irregularities and glucose dysregulation—two significant factors in the development of CVD (12). Research has indicated that elevated glucose levels can hasten endothelial dysfunction, oxidative stress, and inflammation, all of which contribute to atherosclerosis and plaque buildup, thereby raising cardiovascular risk (13, 14). Furthermore, dyslipidemia, especially high cholesterol and low HDL levels, is a recognized risk factor for CVD due to its involvement in plaque formation, impaired reverse cholesterol transport, and heightened vascular inflammation (15, 16). By combining insights from both lipid and glucose metabolism, CHG may deliver a more complete picture of cardiometabolic health (17).

Insulin resistance (IR) and dyslipidemia are closely linked metabolic disorders that contribute significantly to the development of CVD. IR impairs insulin signaling, leading to inefficient glucose uptake and metabolic disturbances, often coexisting with dyslipidemia—characterized by elevated cholesterol levels, low HDL-C, and increased TG (18). Mechanistically, IR stimulates excessive lipolysis in adipose tissue, releasing free fatty acids (FFAs) into circulation. These FFAs are taken up by the liver, promoting the overproduction of very low-density lipoprotein (VLDL), which, in turn, results in increased plasma TG levels, the formation of small, LDL particles, and a decline in HDL-C concentrations. Additionally, hepatic IR exacerbates dyslipidemia by impairing lipid metabolism, fostering an atherogenic profile that elevates CVD risk (19, 20). Considering these metabolic interactions, the CHG index may offer a more comprehensive assessment of cardiometabolic risk. By incorporating glucose levels alongside lipid parameters, CHG provides a broader perspective on metabolic dysfunction, encompassing both dyslipidemia and glucose dysregulation. This innovative approach enhances our ability to evaluate CVD risk more accurately and highlights the importance of considering multiple metabolic pathways rather than isolated lipid markers.

The encouraging results of CHG in this study highlight its potential as a practical and affordable biomarker for assessing CVD risk. Unlike more complicated markers that necessitate advanced testing, CHG is based on routine biochemical parameters that are typically measured in clinical settings. This makes CHG a valuable option for large-scale screening initiatives and primary prevention efforts, especially in areas with limited resources. However, despite CHG showing the highest AUC among the indices examined, its moderate sensitivity and specificity indicate that it should not be relied upon as a sole diagnostic tool. Instead, it may be more effective when used in conjunction with established risk prediction models like the Framingham Risk Score or ASCVD risk calculator to enhance overall risk evaluation. Additionally, longitudinal studies are essential to validate CHG’s predictive ability for future cardiovascular events and its relevance across various populations. Another crucial area for future research is investigating whether CHG can be influenced by lifestyle changes or pharmacological treatments. Given that CHG reflects both lipid and glucose metabolism, it may respond well to dietary adjustments, increased physical activity, and glucose-lowering medications. Understanding how these interventions affect CHG levels could further reinforce its position as a dynamic biomarker for managing cardiovascular risk.

To our knowledge, this is the first study to assess CHG as a new cardiovascular risk marker, combining cholesterol, HDL-C, and glucose into a single measure for evaluating cardiometabolic risk. This research offers important insights into how CHG can predict CVD by comparing its effectiveness with established lipid and metabolic indicators. The large sample size strengthens the reliability of our results, enabling a more robust statistical analysis. Furthermore, the study comprehensively examines at various lipid and glucose-based indices, providing a comparative view of their predictive value for CVD.

This study had several limitations. First, although CHG showed significant links to CVD, further validation in various populations and clinical settings is necessary to generalize our findings. Second, potential confounding factors like dietary habits, physical activity, and medication use were not completely considered, which could affect lipid and glucose metabolism. Lastly, while CHG includes essential metabolic markers, incorporating additional biomarkers, such as those related to inflammation and oxidative stress, could enhance its predictive accuracy for CVD.

## 5. Conclusion

Our findings highlight CHG as a novel and promising biomarker for CVD risk assessment, demonstrating superior predictive ability compared to traditional lipid and metabolic indices. Given its strong association with CVD, CHG has the potential to enhance early risk detection, particularly in individuals with metabolic dysfunction. However, further validation in prospective studies is necessary to establish its clinical utility and integration into existing cardiovascular risk assessment frameworks.

## Declarations

### Ethics approval and consent to participate

All the participants consented to take part in the study by signing written informed consent. The study protocol was reviewed and all methods are approved by the Ethics Committee of Mashhad University of Medical Sciences with approval number IR.MUMS.REC.1386.250. This study was performed in line with the principles of the Declaration of Helsinki. All methods were carried out in accordance with relevant guidelines and regulations.

### Clinical trial number

N/A

### Consent for publication

N/A

### Data availability statement

The datasets used and/or analyzed during the current study available from the corresponding author on reasonable request.

### Author contributions

Amin Mansoori: conception, data analyzing, drafting the article, Mohadeseh Poudineh: drafting the article, Maryam Dianati: drafting the article, Gordon Ferns: revising the article, Mark Ghamsary: data analyzing revising the article, Habibollah Esmaily: corresponding author, Mohsen Moohebati: revising the article, Majid Ghayour-Mobarhan: revising the article.

## Acknowledgments

We would like to thank the MASHAD cohort staff who have participated in running this study.

## Legends

**Figure 1.**
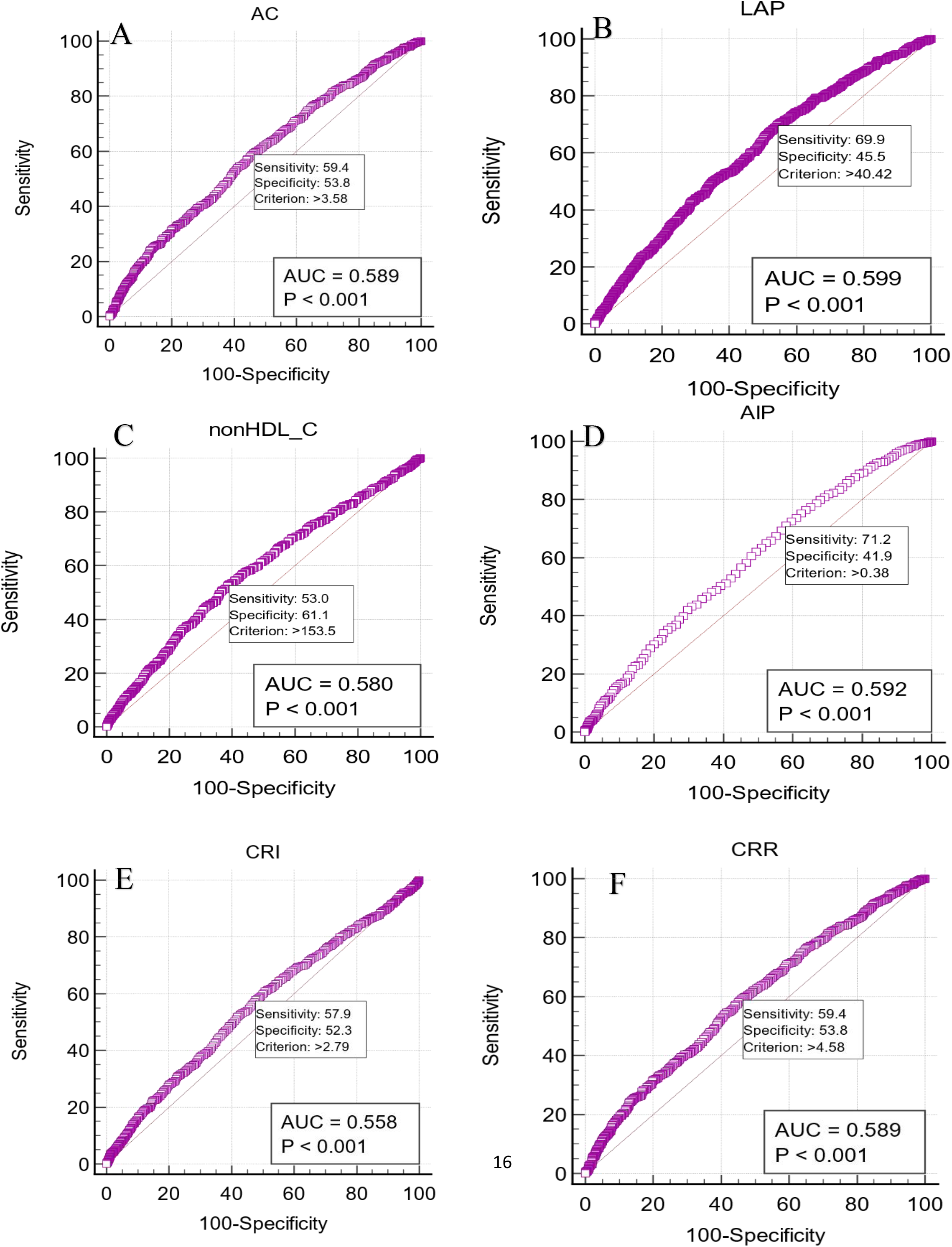
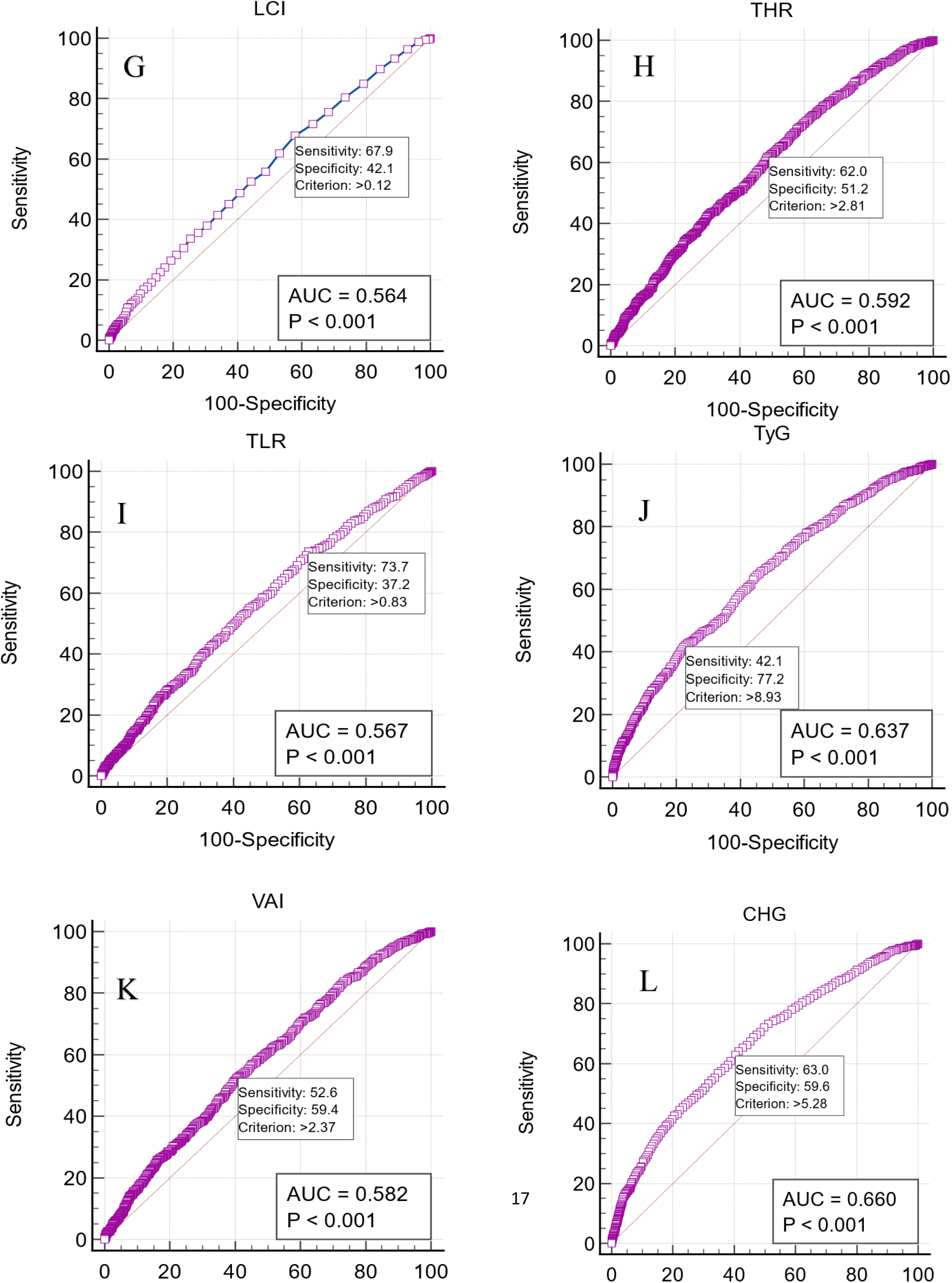
The receiver operating characteristics (ROC) curves for AC (A), LAP (B), nonHDL-C (C), AIP (D), CRI (E), CRR (F), LCI (G), THR (H), TLR (I), TyG (J), VAI (K), CHG (L). p-values in all indices were <0.0001, which shows the differences in area under the ROC curve are statistically significant.

